# Integrated virtual reality and musical biofeedback for intensity-guided training on stationary cycling: a comparative feasibility study

**DOI:** 10.64898/2026.01.14.26343736

**Authors:** Tania Olmo-Fajardo, Prithvi Ravi Kantan, Ana Rojo, Clara B. Sanz-Morère, Erika G. Spaich, Sofia Dahl, Juan C. Moreno

**Affiliations:** BioRobotics Group, Center for Automation and Robotics, Spanish National Research Council, Madrid, Spain; Departamento de Tecnologías de la Información, Escuela Politécnica Superior, Universidad San Pablo-CEU, Madrid, Spain; Department of Architecture, Design and Media Technology, Aalborg University, Denmark; Department of Health Science and Technology, Aalborg University, Denmark

## Abstract

Motor rehabilitation requiring sustained physical exercise faces poor adherence in neurological populations due to insufficient supervision and monotony. While virtual reality and musical biofeedback independently improve engagement and motivation, their comparative and combined impact on intensity control strategies during high-intensity interval training (*HIIT*) remains unexplored.

Thirty healthy adults (16 males, 14 females; mean age 27.5 ± 7.2 years) were sequentially assigned to three feedback modalities (n=10 each) during intensity-guided stationary cycling: visual-only (position-based), musical-only (speed-based), and combined audiovisual (position-based). Participants completed two 9-minute moderate-to-high intensity sessions (Set 1 and Set 2) maintaining pedaling speed within a target speed zone. Performance distinguished control strategy from effectiveness: stability via target zone exits, correction capacity via recovery time and sustained deviations, and overall effectiveness via time in zone. Heart rate (*HR*) assessed physiological intensity; usability and cognitive workload were evaluated via e-Rubric and NASA-TLX.

Distinct regulation strategies emerged. Musical-only showed significantly lower stability (Set 1: 14.52 exits/min vs. 1.48 visual and 1.79 combined; corrected *p <* 0.0167) but superior correction (0.21s recovery vs. 2.48s and 1.06s; *p <* 0.0001) with minimal sustained deviations. Combined feedback achieved highest Set 2 effectiveness (98.13% vs. 95.17% time in zone; corrected *p <* 0.0167) but elevated physical demand (corrected *p <* 0.0167). HR variability was comparable (*p* = 0.85), confirming consistent cardiovascular workload despite differing strategies. Satisfaction was high, with slight preference for musical feedback; cognitive workload did not differ.

Musical biofeedback promotes reactive control with frequent but rapidly corrected oscillations, maintaining physiological safety and engagement. Visual feedback ensures stable target adherence at the cost of compensatory physical effort. Combined modality offered no synergy, increasing demand without improving effectiveness. Findings reveal a trade-off between stability and correction agility, supporting tailored modality selection: musical feedback suits unsupervised rehabilitation prioritizing engagement, rapid error correction, and sustainable effort, while visual feedback suits supervised protocols requiring stable preventive control and precise adherence quantification.

**Author summary:** Many people undergoing neurological rehabilitation struggle to maintain adherence to high-intensity exercise programs, particularly without direct supervision. While virtual reality and musical feedback have shown promise for improving engagement and motivation, we didn’t know which type works best for controlling exercise intensity, or whether combining them would be better.

We tested three feedback systems with 30 healthy adults performing stationary cycling: visual-only, musical-only, and both combined. We measured how well participants stayed within target speed and assessed their experience. Musical feedback prompted frequent but instant adjustments—a reactive strategy that was less physically demanding and most enjoyable. Visual feedback kept participants more precisely in the target zone but required significantly more effort. Surprisingly, combining both didn’t improve performance and instead increased physical demand.

Our results show that different feedback types suit different rehabilitation contexts. Musical feedback may be ideal for unsupervised home-based exercise because it keeps people engaged without requiring exhausting effort. Visual feedback works better when precise control is essential in supervised clinical settings, despite being more demanding. Combining both offers no advantage. These findings help clinicians choose the right feedback approach based on their specific rehabilitation goals.

## Introduction

Effective motor rehabilitation in neurological populations requires sustained, intensive physical exercise to drive functional recovery and neuroplastic adaptation [1–3]. However, this basic requirement faces a critical adherence problem: without constant supervision, patients struggle to maintain prescribed exercise intensity and training frequency, particularly during home-based rehabilitation [4]. This adherence gap is especially problematic given the limited availability of supervised rehabilitation in overloaded healthcare systems, particularly for people affected by stroke and spinal cord injury (*SCI*), where cardiovascular complications are the leading cause of long-term morbidity and mortality [5–7]. The challenge is twofold: rehabilitation protocols must be sufficiently intense to yield clinical benefit and engaging enough to sustain long-term participation when direct supervision is unavailable.

Stationary cycling represents an accessible, safe, and biomechanically relevant modality for lower limb rehabilitation. Its cyclic movement patterns share kinematic similarities with gait, require considerable joint range of motion, and support muscle synergy reorganization underlying motor learning [8–13]. High-intensity interval training (*HIIT*), involving repeated short bursts of high-intensity exercise alternating with active recovery periods, has emerged as a particularly promising approach for aerobic training, achieving superior cardiovascular and metabolic benefits compared to continuous moderate-intensity exercise in shorter durations [14–16]. In neurorehabilitation, HIIT cycling protocols can help to improve cardiorespiratory fitness, muscular strength, and motor function in stroke patients while potentially driving greater neuroplastic changes crucial for long-term recovery [17–22]. The efficacy of HIIT depends on precise intensity control to ensure patients reach and maintain target cardiovascular zones, based on heart rate (*HR*), while avoiding dangerous overshoots [23–25]. While automated HR-controlled ergometer systems have been developed to manage this [26, 27], HR responds with inherent physiological delay to changes in physical effort [28, 29]. This creates a challenge in feedback control strategies: patients must regulate their pedaling speed (an immediately controllable parameter) to achieve desired HR targets (a delayed physiological response). This is particularly difficult to implement without constant supervision in clinical settings where therapists must oversee multiple patients simultaneously [30]. To address this issue, we propose providing real-time feedback on immediately controllable parameters (position, speed) to enable patient self-regulation, keeping the user “in-the-loop” while HR is monitored for safety.

Two technology-mediated feedback approaches have independently demonstrated potential for addressing these challenges by providing real-time guidance on pedaling speed. Virtual reality (VR) offers immersive, interactive environments that reduce pain perception during high-intensity exercise and increase motivation compared to conventional methods [31–33]. VR-based cycling interventions have shown clinical efficacy in neurological populations: Yin et al. demonstrated that VR cycling with force plate feedback significantly improved bilateral lower limb strength and standing balance in post-stroke patients [34], while Yang et al. found that VR cycling with biofeedback enhanced walking endurance, speed, and reduced muscle spasticity compared to traditional training [35]. Systems like PedaleoVR and iCycle have confirmed feasibility and high usability in patients with ataxia and spinal cord injury [36, 37], and recent paradigms such as Buddy Biking [38] have implemented VR cycling in social contexts. The visual modality provides rich spatial information enabling anticipatory control through continuous feedback about position relative to targets.

Musical biofeedback, on the other hand, takes advantage of the auditory system’s high temporal resolution and strong neural connectivity to motor areas [39, 40]. The auditory system’s superior temporal processing makes it particularly suited for providing immediate, reactive feedback on movement parameters. Music-based biomechanical feedback systems have successfully guided gait and balance training [41–43], and studies confirm that music reduces perceived exertion, increases enjoyment, and helps regulate movement parameters like cadence [44–46]. By modulating musical features (tempo, volume, pitch) in real-time based on movement speed or other kinematic parameters, musical feedback can serve as an engaging extrinsic cue that supports self-regulation without requiring constant visual attention [42].

Multisensory integration theory suggests that combining visual and auditory modalities could enhance motor learning and control by making behaviorally relevant events more noticeable through temporal coincidence and cross-modal reinforcement. However, this benefit relies on spatiotemporal congruency between stimuli to approximate real-world sensory interactions [47, 48]. Recent clinical trials support this premise in gait rehabilitation: combined visual feedback and rhythmic auditory cues during treadmill training significantly improved gait symmetry and balance in chronic stroke patients [49], while a multi-center trial confirmed that audiovisual cueing enhanced lower limb sensorimotor recovery in hemiplegic patients [50]. However, multisensory benefits are not guaranteed: psychophysical research demonstrates that visual and auditory signals do not always integrate automatically into a single, more efficient percept, particularly when modalities convey conflicting or redundant information [51]. Strong intramodal interference can limit or even negate potential benefits of multimodal feedback. While these studies demonstrate promise in gait training, they leave fundamental questions unanswered regarding feedback selection and design for intensity-guided cycling exercise. Despite growing use of virtual technologies combined with music in neurorehabilitation for walking and coordination exercises [52], their application to cycling has not been reported.

Significant knowledge gaps prevent evidence-based implementation of multimodal feedback for intensity-guided cycling exercise. First, to the best of our knowledge, no study has directly compared visual and musical feedback modalities within the same cycling task using equivalent performance metrics, making it impossible to determine their relative advantages for intensity control. Second, whether combining these modalities in cycling produces synergistic benefits or cognitive interference remains unknown, as existing multimodal studies focus on gait rather than cycling and have not systematically isolated the contribution of each sensory channel. Third, optimal feedback design principles for intensity-guided cycling remain unestablished, specifically regarding the critical trade-offs between control strategies (stability vs. correction agility), overall effectiveness, physiological load, and user experience. Finally, the application of combined VR and musical feedback to HIIT-structured cycling represents a significant translational gap given the clinical importance of precise intensity control in interval training protocols.

To address these gaps, we developed an integrated VR and musical biofeedback platform for HIIT-structured stationary cycling and conducted a comparative feasibility study in healthy adults. Our research questions were: (RQ1) What are the performance trade-offs between these modalities in terms of control strategy (stability vs. correction agility) and overall effectiveness in maintaining target exercise intensity? (RQ2) Does combined feedback produce synergistic benefits or impose cognitive penalties relative to unimodal feedback? (RQ3) How do visual-only, musical-only, and combined audiovisual feedback modalities compare in usability and user satisfaction when applied to intensity-guided cycling? (RQ4) Do these modalities maintain comparable physiological load as measured by cardiovascular response?

Based on multisensory integration theory and recent clinical evidence showing benefits of combined audiovisual cueing in gait rehabilitation [49, 50], we hypothesized that combined feedback would achieve superior adherence to target pedaling speed and higher user satisfaction compared to either unimodal condition. Furthermore, we expected music-only feedback to demonstrate higher satisfaction than visual-only feedback due to music’s capacity to act as a natural reward stimulus, triggering dopaminergic release that regulates mood and reinforces the intrinsic motivation to exercise [53, 54]. This proof-of-concept study in healthy adults establishes an empirical foundation for evidence-based selection of feedback modalities in future clinical trials with neurological populations, where adherence and precise intensity control are essential rehabilitation outcomes.

## Materials and methods

### System description and interaction design

In the initial development phase of the combined modality, two independent components for VR and musical feedback (linked to cumulative and instantaneous pedaling speed, respectively) were coupled and evaluated in a pilot study with 11 participants [55, 56]. Following an iterative optimization process, a second development cycle led to a robust integration of both components and to a refined combined feedback design focused on cumulative pedaling speed. This process resulted in an individualized feedback system that served as the foundation of the system employed in the present study.

#### Design rationale and multisensory integration principles

The multimodal feedback system was designed to communicate pedaling speed through congruent visual and auditory channels, employing principles of multisensory integration [47, 48] and crossmodal congruency [55]. The design exploited the natural relationship between cycling speed and forward displacement to create an intuitive virtual environment where users control a bicycle navigating a scenic track. Visual feedback offered precise spatial information and goal-oriented guidance, while musical feedback provided continuous temporal cues that could be processed with minimal attentional demand [46]. This combination aimed to enhance motivation through the known benefits of music during exercise [44, 45] while maintaining precise intensity control through visual goal-directed behavior. The feedback system provided guidance on immediately controllable pedaling parameters (position or speed) rather than delayed heart rate response. While HR was monitored to validate physiological safety, it was not incorporated into the real-time feedback loop. This user-in-the-loop design prioritizes engagement and system simplicity while ensuring appropriate cardiovascular intensity through offline verification.

#### Visual feedback design

The virtual environment presented a first-person perspective of cycling along a scenic track rendered in Unity3D (v.2021.3.16). A virtual rabbit moving ahead of the user’s bicycle (Fig 2E) represented the target speed, exploiting the natural association between pedaling effort and translational speed in real-world cycling [30]. The user’s goal was to maintain a constant distance from the rabbit by matching target pedaling speed. To support this goal, the system relied primarily on natural visual perception to provide continuous feedback. The distance between the bicycle and the rabbit indicated cumulative deviation from the target speed, where falling behind expressed sustained under-target performance and overtaking indicated sustained over-target performance. This distance-based representation aligns with established principles of continuous visual feedback for motor control [57]. Simultaneously, optic flow reflected instantaneous pedaling speed, providing peripheral awareness of the current effort level consistent with ecological perception principles [58]. Complementing these natural cues, the sky color served as an extra, discrete signal to clarify performance zones. Based on focus group discussions and gaming conventions [59], the sky appeared light blue within the target range, shifting to red when the user was too slow and yellow when too fast. This ensured that significant deviations were immediately noticeable without interfering with the primary tracking task.

#### Musical feedback design

Musical feedback was implemented through the manipulation of the playback rate of pre-selected music tracks, which simultaneously altered both tempo and pitch. This design exploited the auditory system’s high temporal resolution for precise movement guidance [39] and the well-established association between musical tempo and the general construct of speed to minimize cognitive load [40]. The control variable for this feedback was adapted based on the feedback modality. In the audiovisual condition, the playback rate was coupled to the distance between the bicycle and the rabbit such that lagging too far behind slowed the music whilst overtaking the rabbit sped it up beyond its nominal rate (see S1 Video). This matched the visual representation to maintain crossmodal congruency and support multisensory integration [48], creating redundant information across modalities. In the musical-only condition, however, the playback rate was coupled to instantaneous pedaling speed. This adaptation was necessary because cumulative position lacks an intuitive auditory equivalent without a visible spatial frame of reference (the rabbit), whereas speed has a direct perceptual correlate in temporal rate. In this condition, normal-sounding music indicated performance within the target speed zone. Playback rate modulation was constrained to ±20% of normal tempo to maintain musical coherence and avoid excessive distortion.

#### System architecture and implementation

The integrated platform comprised two networked applications running on Windows PCs connected via local wireless network (WLAN): a C++ based application for sensor processing (made using the JUCE environment) and audio control, and a Unity3D application for visual rendering (Fig 1). Communication between applications used Open Sound Control (OSC) protocol. An M5Stack Core2 inertial measurement unit (IMU), attached to the participant’s lateral thigh (Fig 2B), transmitted 3D accelerometer and gyroscope data wirelessly at 100 Hz to the C++ application. All cycling was performed using a MOTOmed Viva2 ergometer (RECK-Technik GmbH & Co., Germany; Fig 2A), but the protocol is generalizable to any stationary bicycle as the control was based on the external thigh-mounted sensor rather than ergometer-internal measurements.

**Fig 1.**
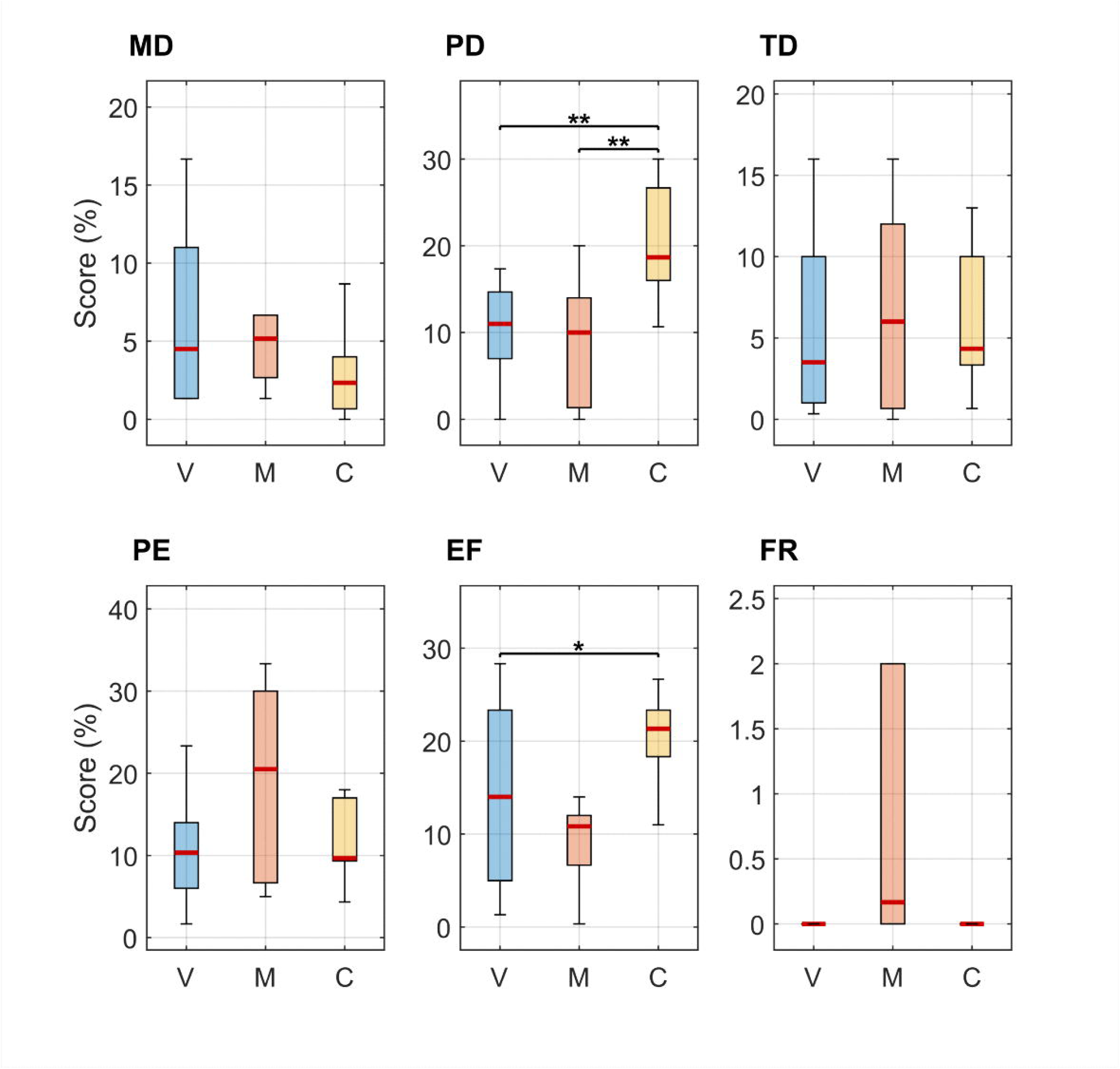
Block diagram illustrating data flow from physical measurement to user feedback presentation. The M5 Core2 sensor unit attached to the user’s thigh sends data (via WiFi/OSC) to the C++ Application, which processes thigh rotation velocity and normalization algorithms to determine real-time speed, position, and target status (highlighted in yellow). These computed variables drive both sound rendering (musical feedback) and the Unity application (VR feedback).

**Fig 2.**
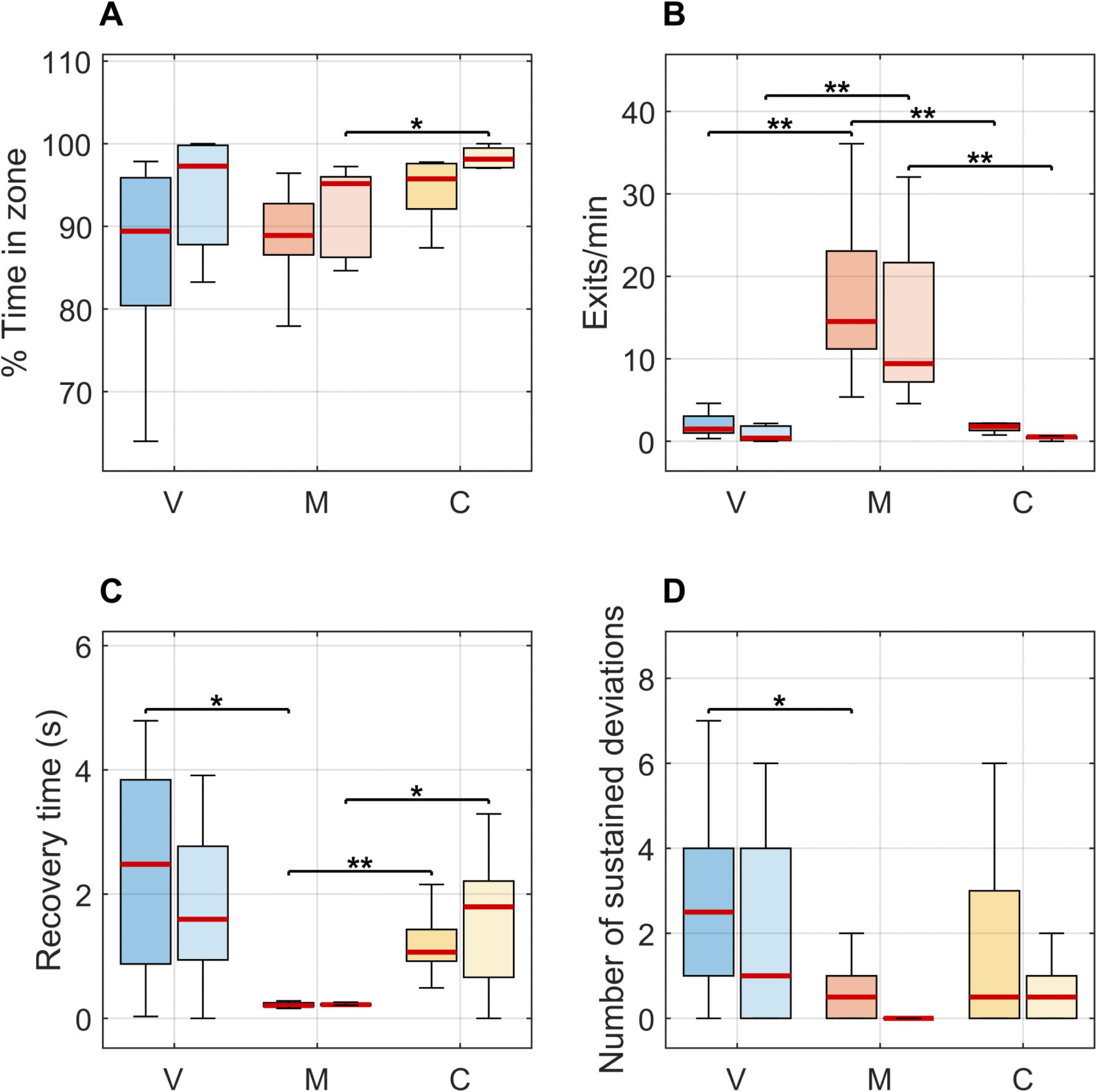
Overview of the cycling feedback platform and its components. Schematic representation of healthy volunteer cycling on a MOTOmed Viva2 ergometer (A) while wearing an M5Core2 inertial sensor on the thigh (B) and headphones for musical feedback (C). The platform comprises several integrated components: the feedback system user interface for experimenter control (D), the immersive virtual reality environment displaying the track and target rabbit (E), and the heart rate monitoring display showing real-time cardiovascular data (F).

Pedaling speed (quantified as smoothed thigh rotation speed) was estimated from sagittal-plane angular velocity using the following signal processing pipeline: (1) half-wave rectification to isolate forward rotation phases, (2) low-pass filtering (2nd-order Butterworth, cutoff 2 Hz) to remove high-frequency noise, and (3) envelope detection (attack time 200 ms, release time 200 ms) as implemented in [60, Chapter 13] to obtain a smoothed speed signal. The C++ application implemented individualized calibration by recording desired pedaling speed during a baseline period, establishing each participant’s reference value that was used to scale all subsequently computed speed values to fit a normalized 0-1 range.

The core control algorithm computed three variables every 10 ms: (1) user’s instantaneous pedaling speed as explained above, (2) cumulative position of the virtual bicycle (pedaling speed integrated over time), and (3) cumulative position of the rabbit (target/rabbit speed integrated over time). The rabbit’s speed could graphically be programmed to vary throughout a session, enabling the implementation of structured speed intervals corresponding to different physiological training intensities. Target speed zones were defined relative to the baseline reference speed established during calibration. For position-based feedback (visual-only and combined audiovisual), bicycle and rabbit positions were compared to determine cumulative deviation, classified into three zones: too fast (any position ahead of target, *>* 0 virtual units), within-target range (0 to 0.8 virtual units behind target), and too slow (more than 0.8 virtual units behind target). For speed-based feedback (musical-only), instantaneous speed was compared to target speed with an acceptable range of ±0.1 virtual units per second. These thresholds were empirically determined through pilot testing to approximately balance deviation penalties across modalities.

The computed control variable (position deviation or speed deviation) was transmitted via OSC to drive auditory and visual feedback. For auditory feedback (Fig 2B), the C++ application included an integrated music player that loaded user-selected tracks and directly manipulated playback rate in real-time based on the deviation signal. For visual feedback, computed speeds of bicycle and rabbit were transmitted via OSC to Unity, which updated the 3D environment (Fig 2E) at *>* 100 frames per second on a high-performance gaming PC with dedicated graphics card, rendering bicycle and rabbit positions and updating sky color based on deviation from target. The C++ application provided an experimenter interface for session control, including sensor calibration tools, real-time monitoring, programmable target speed profiles, music selection, and adjustable difficulty parameters (Fig 2D). At session completion, the application generated summary reports and logged all sensor and computed variables at 100 Hz for offline analysis.

### Experimental design and protocol

#### Ethics and study design

This study received ethical approval from the Spanish National Research Council (CSIC) Ethical Committee (Reference: 306/2023). The recruitment period for this study spanned from 24/06/2024 to 16/10/2024. All participants provided written informed consent prior to participation. The study employed a between-subjects design with three parallel groups corresponding to different feedback modalities.

#### Participants

A convenience sample of 30 healthy adults participated (16 males, 14 females; mean age 27.50 ± 7.21 years, median 25 years, range 20-46 years). Participants were consecutively assigned to one of three intervention groups by order of arrival to achieve equal group sizes (n=10 per group). Group demographics were balanced: visual-only (6 males, 4 females; mean age 29.40 ± 8.17 years), musical-only (5 males, 5 females; mean age 29.00 ± 8.52 years), and combined audiovisual (5 males, 5 females; mean age 24.10 ± 3.00 years).

Inclusion criteria required healthy individuals aged 18-65 years of either sex with no motor impairments affecting lower limb function. Exclusion criteria were uncorrected visual or auditory deficits, medical contraindications to physical activity (e.g., cardiovascular conditions, recent orthopedic surgery), acute illness at time of testing, or direct involvement in the research project.

#### Experimental procedure

Upon arrival, participants were instrumented and positioned on the ergometer. Before data collection began, they were provided with a detailed explanation of the procedure.

#### Baseline calibration and target intensity determination

Participants underwent initial calibration to determine individualized baseline reference speed corresponding to moderate-to-high cardiovascular effort. This baseline speed was defined as the pedaling speed required to reach and sustain 70% of age-predicted maximum heart rate (HRmax), calculated using sex-specific formulas: Tanaka equation for males (HRmax = 208.75 - 0.73 × age) [61] and Gulati equation for females (HRmax = 206 - 0.88 × age) [62]. This threshold represents the moderate-to-high intensity training zone based on the Karvonen method [63]. Heart rate was monitored at 1 Hz using a Polar H10 chest strap sensor (Fig 2F) throughout all sessions to validate physiological intensity, though HR data was not provided as real-time feedback to participants. This calibration phase lasted 5-10 minutes.

#### Exercise protocol

Following a rest period of up to 10 minutes to allow HR return to baseline, participants completed two 9-minute structured cycling sessions with varying target speeds, inspired by high-intensity interval training (HIIT) principles. **Set 1** consisted of 3 minutes at baseline reference speed, 3 minutes at +15% baseline speed, and 3 minutes at baseline speed. Subsequently, **Set 2** comprised 3 minutes at baseline speed, 3 minutes at -15% baseline speed, and 3 minutes at baseline speed.

A minimum 10-minute rest period separated the two sets to allow HR recovery to within 10 bpm of baseline. Set order was fixed (Set 1 followed by Set 2) to prioritize participant safety by placing the most physiologically demanding set (+15%) when participants were fully rested, preventing accumulated fatigue from compromising task completion. All 30 participants completed both sets without adverse events.

#### Post-session assessment

Immediately after finishing both exercise sets, participants completed two standardized questionnaires assessing overall system usability and cognitive workload across the entire session (both sets). Perceived exertion was assessed using the Borg CR10 scale [64] before and after each individual set to monitor fatigue levels and verify participants’ readiness to proceed.

### Outcome measures and data collection

#### Primary outcomes: performance metrics

Pedaling speed data from the IMU sensor were used to compute feedback-agnostic metrics enabling direct comparison across different feedback modalities. These performance metrics distinguished between overall effectiveness and control strategies used to maintain the target speed.

To assess overall effectiveness, we calculated the *percentage of time within target speed zone (% time in zone)*. This metric represents the proportion of session duration during which pedaling speed remained within the predefined target speed zone, quantifying sustained adherence to the prescribed training intensity.

Furthermore, control stability was evaluated using the *rate of exits from target speed zone (exits/min)*. Defined as the median number of exits from the target zone per minute, high rate of zone exits indicate unstable control characterized by frequent oscillations around the target, which could potentially compromise training zone specificity in interval protocols.

Correction capacity was assessed through two complementary metrics. First, *recovery time (seconds)* measured the median duration of excursions outside the target zone, where shorter times indicate more effective real-time error correction. Second, the *number of sustained deviations* counted periods where pedaling speed remained outside the target speed zone for more than 5 consecutive seconds. This safety-relevant metric identifies significant sustained deviations that might compromise physiological training intensity or pose risks in clinical populations.

These metrics were selected because they apply uniformly across position and speed-based feedback systems. They directly reflect clinically relevant training outcomes by characterizing distinct aspects of user performance (effectiveness, stability, and correction capacity) while avoiding the methodological limitations of comparing error magnitudes across incompatible target representations.

### Secondary outcomes

#### Physiological monitoring

Heart rate was recorded throughout all sessions using a Polar H10 chest strap sensor to check that participants maintained appropriate cardiovascular intensity (Fig 2F). HR data served as an independent physiological measure to confirm that differences in pedaling control strategies did not result in different cardiovascular workloads across feedback modalities.

#### Usability assessment

System usability and user satisfaction were evaluated using a questionnaire based on the e-Rubric methodology [65], which assesses technical usability dimensions including accessibility, ease of handling, and interface design. The questionnaire comprised 21 applicable items rated on a Likert scale from 1 to 5 (one item removed as non-applicable; see S1 Appendix), with alternating direct and reverse-worded statements to reduce response bias. The instrument demonstrated high internal consistency (Cronbach’s alpha = 0.889). Items were grouped into four dimensions following the USE framework: *Usefulness* (2 items), *Ease of Use* (7 items), *Ease of Learning* (4 items), and *Satisfaction* (8 items).

The e-Rubric questionnaire was selected over more general instruments such as the System Usability Scale (SUS) [66] or the USE questionnaire [67] because it provides detailed evaluation of interactive feedback systems while maintaining necessary conciseness, offering an optimal balance between comprehensiveness and participant burden.

#### Cognitive workload assessment

Cognitive workload was measured using the NASA Task Load Index (NASA-TLX) [68] with the weighted procedure. Participants rated six dimensions: *Mental Demand (MD)*, *Physical Demand (PD)*, *Temporal Demand (TD)*, *Performance (PE)*, *Effort (EF)*, and *Frustration (FR)*. Weighted scores were normalized to 0-100% scale for interpretability, with ranges defined as low (0-33%), medium (34-66%), and high (67-100%) workload.

### Statistical analysis

For each outcome variable, normality was assessed using Shapiro-Wilk tests (*α* = 0.05) and homogeneity of variances using Bartlett’s test (normally distributed data) or Levene’s test (non-normally distributed data).

To test for main effects of feedback modality on outcome measures, between-group comparisons used:

- One-way ANOVA followed by Tukey-Kramer post-hoc tests (normal distribution, homogeneous variances)
- Welch ANOVA followed by Games-Howell post-hoc tests (normal distribution, heterogeneous variances)
- Kruskal-Wallis test followed by pairwise Mann-Whitney U tests (non-normal distribution)

Given the small sample size and non-normal distributions in several outcomes, all descriptive statistics are reported as median [IQR]. Statistical significance was set at *α* = 0.05 for omnibus tests and *α* = 0.0167 for post-hoc pairwise comparisons (Bonferroni-adjusted for three comparisons: visual vs. musical, visual vs. combined, musical vs. combined). Effect sizes (*η*^2^) were interpreted as small (0.01-0.06), medium (0.06-0.14), and large (*>* 0.14) [69]. All analyses were performed using MATLAB R2024a, incorporating functions from the Statistics and Machine Learning Toolbox.

## Results

All participants (n=30) completed the calibration phase and both exercise sets without adverse events, technical failures, or dropouts. The following sections detail the effects of feedback modality on performance, physiological response, cognitive workload, and user experience.

### Performance metrics: effectiveness and control strategies

All feedback modalities successfully facilitated adherence to the prescribed speed targets. Fig 3 presents the key performance metrics for both exercise sets, illustrating the distinct control strategies employed across feedback modalities. Detailed performance metrics across both exercise sets are provided in S1 Table, S2 Table, S3 Table and S4 Table. The median speed deviation relative to target zone thresholds for each group is provided in S1 Fig.

**Fig 3.**
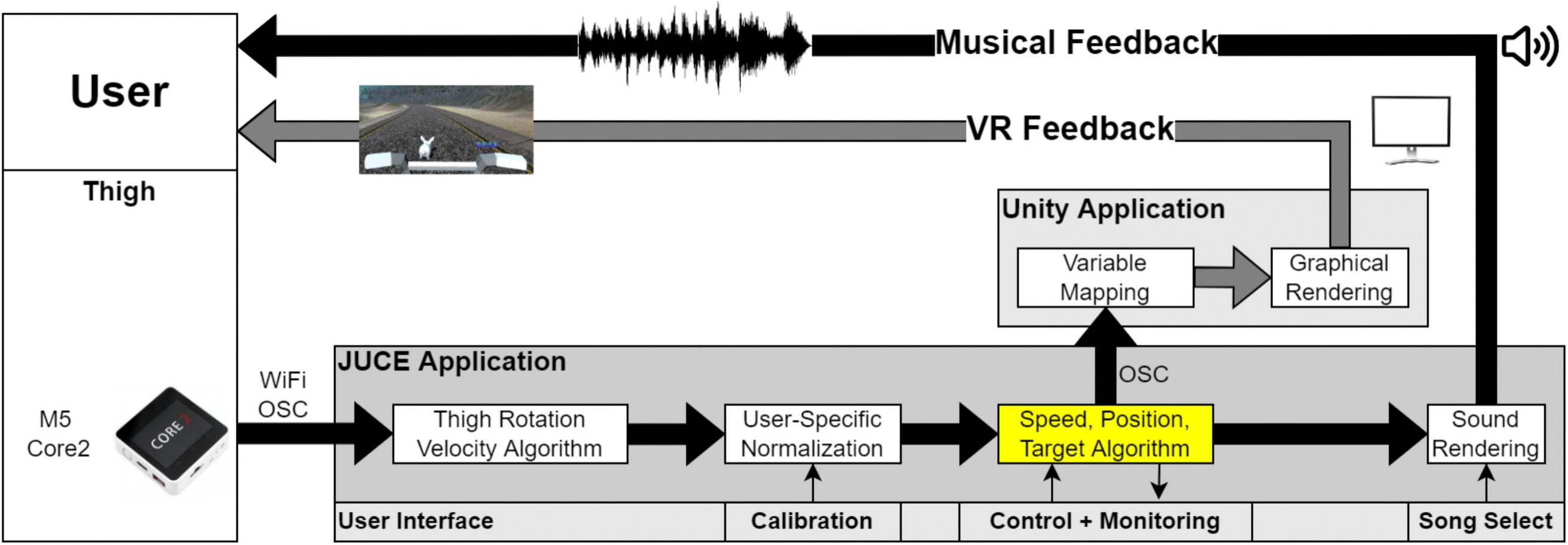
Performance metrics across feedback modalities. Comparison of cycling performance measures for visual-only (V, blue), musical-only (M, orange), and combined audiovisual (C, yellow) feedback conditions during both Set 1 (darker) and Set 2 (lighter). The red line within each box indicates the median value, the box edges represent the interquartile range (25th to 75th percentiles), and the whiskers extend to the most extreme data points not considered outliers. Asterisks denote statistically significant pairwise differences following Bonferroni correction: \**p <* 0.0167, \*\**p <* 0.001. **(A)** Percentage of time spent within the target speed zone. **(B)** Rate of exits from the target zone (exits per minute) **(C)** Recovery time (seconds) representing the median duration of excursions outside the target zone. **(D)** Number of sustained deviations (periods exceeding 5 consecutive seconds outside target zone).

All three modalities achieved high adherence to target speed zones, with percentage time in zone ranging from 89% to 98% across groups and sets (Fig 3A). The feedback modality affected adherence to target speed zones for Set 1 (Welch ANOVA: *p* = 0.0335, *η*^2^ = 0.155), though post-hoc analysis did not show any pairwise comparison meeting the Bonferroni-adjusted significance threshold (*α_adj_*= 0.0167). All three groups achieved high median adherence rates (visual: 89.42% [80.41–95.88], musical: 88.90% [86.55–92.75], combined: 95.75% [92.10–97.60]). For Set 2, the feedback modality significantly affected zone adherence (Welch ANOVA: *p* = 0.0164, *η*^2^ = 0.190), with post-hoc tests revealing that the combined audiovisual condition achieved significantly higher adherence compared to the musical condition (combined: 98.13% [97.10–99.48] vs. musical: 95.17% [86.26–95.99]; *p* = 0.0077).

While overall effectiveness was comparable, the control strategy employed differed significantly. The feedback modality significantly affected the *rate of zone exits* in both Set 1 (Welch ANOVA: *p* = 0.0006, *η*^2^ = 0.661) and Set 2 (Kruskal-Wallis: *p* = 0.0001, *η*^2^ = 0.573) (Fig 3B). In both sets, the musical group showed a significantly higher frequency of exits per minute compared to both the visual and combined conditions (*p <* 0.0005 for all four comparisons). In Set 1, musical feedback showed a median of 14.52 [11.19–23.07] exits/min, approximately 10-fold higher than visual (1.48 [1.00–3.05]) and 8-fold higher than combined (1.79 [1.31–2.18]). This pattern persisted in Set 2, with musical exhibiting 9.41 [7.19–21.67] exits/min compared to visual (0.38 [0.11–1.84]) and combined (0.54 [0.32–0.55]).

Correction capacity, measured by *recovery time*, was also significantly affected by feedback modality for both Set 1 (Welch ANOVA: *p <* 0.0001, *η*^2^ = 0.479) and Set 2 (Welch ANOVA: *p* = 0.0009, *η*^2^ = 0.266) (Fig 3C). In Set 1, musical feedback exhibited significantly faster recovery times (0.21s [0.19–0.25]), approximately 10-fold shorter than visual (2.48s [0.87–3.84]; *p* = 0.0022) and combined (1.06s [0.92–1.43]; *p <* 0.0001). In Set 2, musical (0.22s [0.21–0.23]) remained significantly faster than combined (1.80s [0.66–2.21]) (*p* = 0.0014).

The feedback modality affected the number of *sustained departures* from target speed (Fig 3D). The number of sustained deviations (periods outside the zone for *>* 5 s) showed significant overall differences in both Set 1 (Kruskal-Wallis: *p* = 0.0422, *η*^2^ = 0.173) and Set 2 (Kruskal-Wallis: *p* = 0.0369, *η*^2^ = 0.249). In Set 1, post-hoc analysis indicated that musical feedback resulted in significantly fewer sustained deviations than visual feedback (*p* = 0.0109) (musical: 0.50 [0.00–1.00] vs. visual: 2.50 [1.00–4.00]). In Set 2, despite the overall significance, no pairwise comparison reached the adjusted threshold (*α_adj_* = 0.0167). The musical group had a median of 0.00 [0.00–0.00] sustained deviations.

### Physiological response

Heart rate variability, quantified as the coefficient of variation of HR (CV-HR), showed no significant effect of feedback modality across the three conditions (ANOVA: *p* = 0.8518, *η*^2^ = 0.006) (see S2 Fig). Furthermore, analysis of heart rate trajectories throughout the exercise sessions confirmed that all three feedback modalities maintained participants within the prescribed cardiovascular training zones (above 70% of age-predicted HRmax) with similar temporal profiles (see S1 Fig).

### Cognitive workload

Cognitive workload scores for all NASA-TLX subscales and the weighted total score are shown in S5 Table. Overall cognitive workload (*TLX_total_*) showed no significant difference of feedback modality (ANOVA: *p* = 0.2450), with median values indicating a moderate load for all conditions: visual (54.8 [48.3–58.7]), musical (51.2 [42.0–70.0]), and combined (64.5 [51.3–69.7]) (Fig 4 and S3 Fig). However, analysis of the weighted dimensions (see S6 Table) revealed significant effects on *Physical Demand (PD)* and *Effort (EF)*.

**Fig 4.**
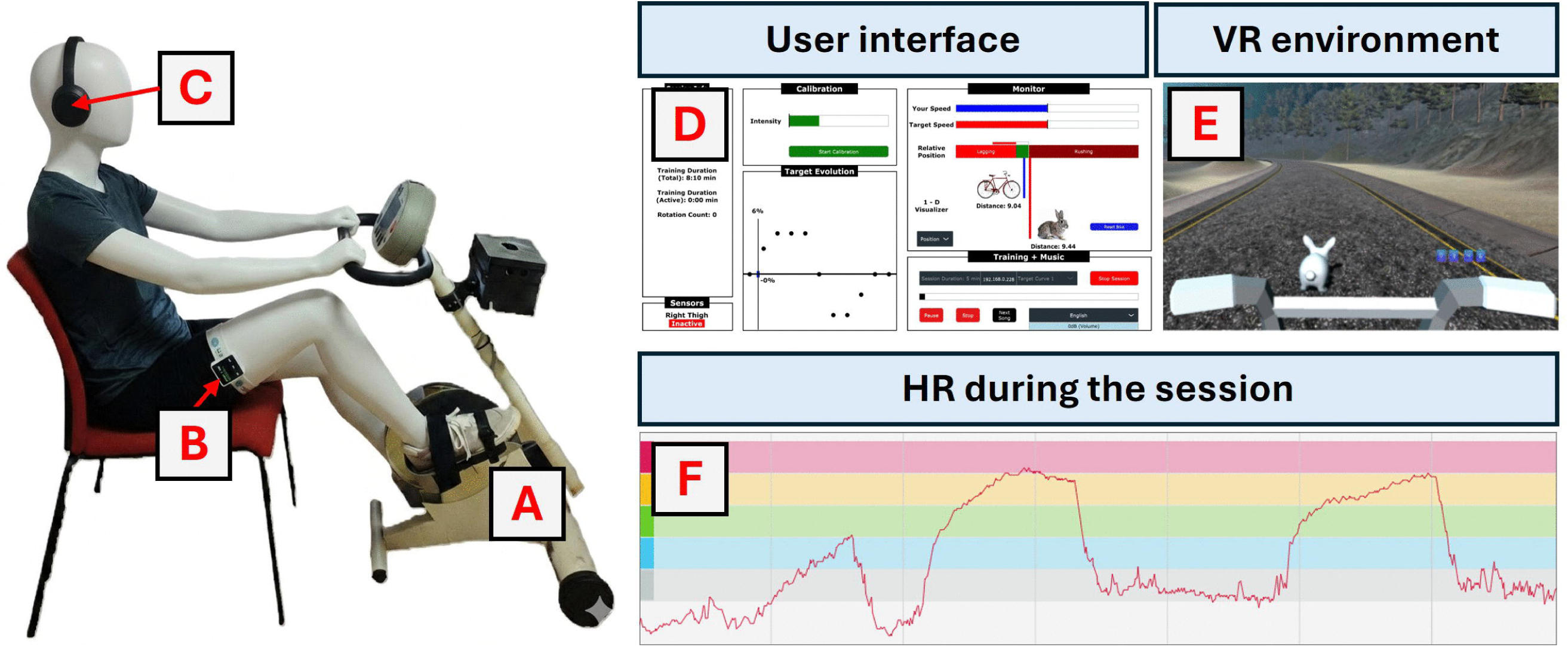
NASA-TLX weighted scores by dimension. Distribution of scores (0–100%) for the six subscales of the *NASA-TLX* questionnaire across visual-only (V, blue), musical-only (M, orange), and combined audiovisual (C, yellow) feedback conditions. The red line within each box indicates the median score, the box edges represent the interquartile range (25*^th^* to 75*^th^* percentiles), and the whiskers extend to the most extreme reported scores not considered outliers. Asterisks denote statistically significant pairwise differences: \**p <* 0.0167, \*\**p <* 0.01. *Abbreviations:* MD: Mental Demand; PD: Physical Demand; TD: Temporal Demand; PE: Performance; EF: Effort; FR: Frustration Level.

The feedback modality significantly affected perceived physical demand (ANOVA: *p* = 0.0016, *η*^2^ = 0.378)(Fig 4PD). Post-hoc analysis (Tukey-Kramer) indicated that the combined group reported significantly higher physical demand (PD) (18.7% [16.0–26.7]) compared to both the musical (10.0% [1.3–14.0]; *p* = 0.0090) and visual (11.0% [7.0–14.7]; *p* = 0.0025) groups. The perceived physical demand for the combined feedback condition was approximately double that of the musical feedback condition. On the 0-100% scale, these median values indicate low-to-moderate physical demand across all conditions, with the combined condition reaching the upper range of moderate demand.

A significant difference was also observed for perceived effort (EF) (ANOVA: *p* = 0.0189, *η*^2^ = 0.255) (Fig 4EF). Post-hoc tests indicated that the combined group required significantly higher effort (21.3 [18.3–23.3]) compared to the visual group (14.0 [5.0–23.3]; *p* = 0.0147). These median values indicate low-to-moderate effort across all conditions.

No significant effects of feedback modality were found in the remaining NASA-TLX dimensions: mental demand (MD) (Kruskal-Wallis: *p* = 0.1486), temporal demand (TD) (ANOVA: *p* = 0.8039), performance (PE) (ANOVA: *p* = 0.1164), or frustration (FR) (Kruskal-Wallis: *p* = 0.1725). Importantly, median scores for mental demand, temporal demand, and frustration remained low across all modalities (all below 7% on the 0-100% scale), suggesting that none of the feedback conditions imposed excessive cognitive workload. Regarding the performance dimension (where higher values indicate better perceived performance), while differences were not significant, the musical group reported a noticeably higher median score (20.5% [6.7–30.0]) compared to visual (10.3% [6.0–14.0]) and combined (9.7% [9.3–17.0]).

### User experience

All three feedback modalities achieved high usability scores across e-Rubric dimensions, with median scores generally exceeding 4.0 on a 5-point scale, indicating strong perceived feasibility for all tested conditions (Fig 5 and S4 Fig). The feedback modality did not significantly affect any usability dimension (see S7 Table and S8 Table): Usefulness (Kruskal-Wallis: *p* = 0.0789), Ease of Use (ANOVA: *p* = 0.3739), Ease of Learning (ANOVA: *p* = 0.7868), Satisfaction (ANOVA: *p* = 0.0889), or Overall Usability (ANOVA: *p* = 0.0698).

**Fig 5.**
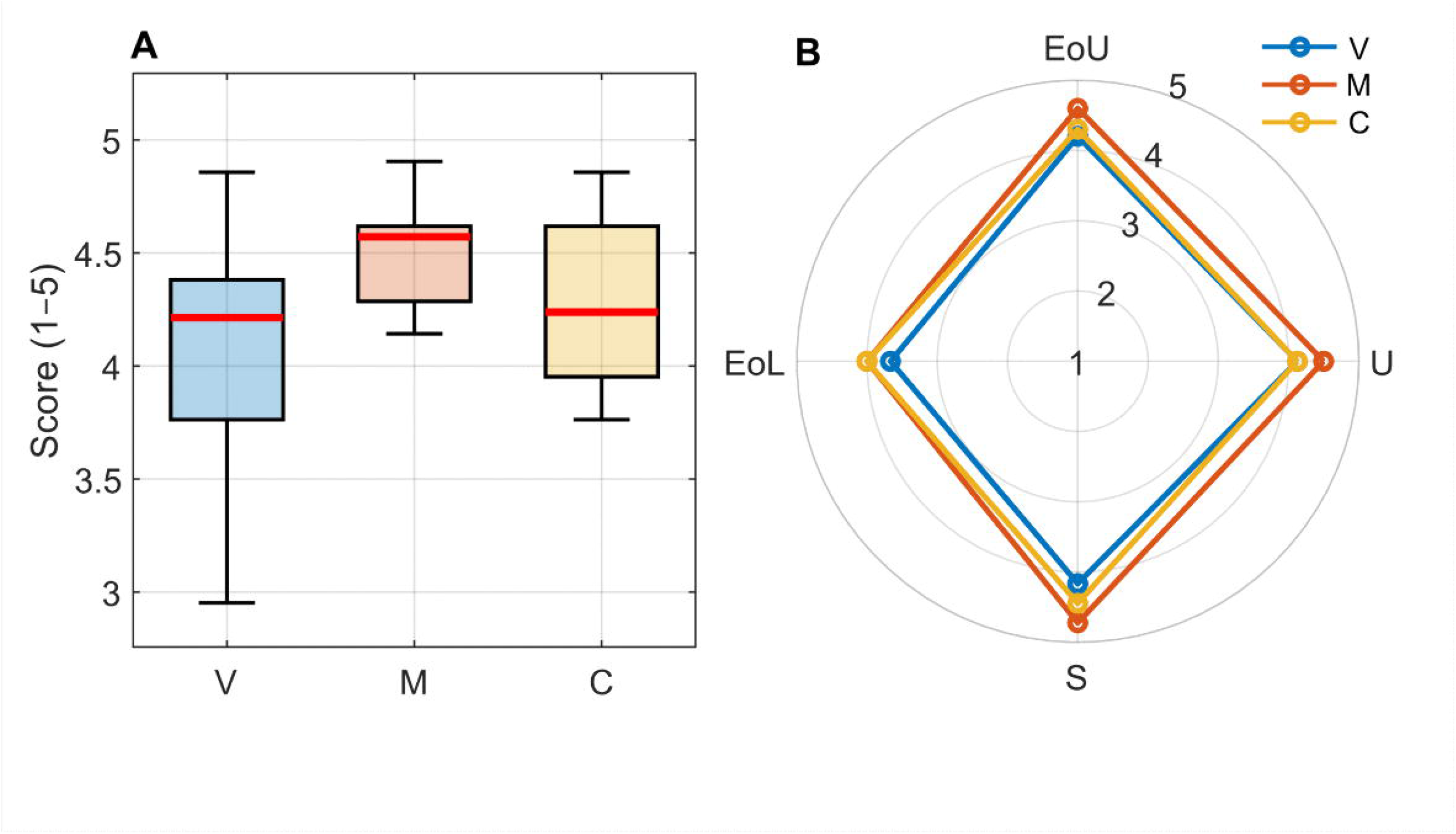
Overall user experience scores and radar chart showing the different components. **(A)** Overall user experience scores (1–5 Likert scale) reported using the e-Rubric questionnaire for the three feedback modalities: visual-only (V, blue), musical-only (M, red), and combined audiovisual (C, yellow). The red line within each box indicates the median score, the box edges represent the interquartile range (25th to 75th percentiles), and the whiskers extend to the most extreme reported scores not considered outliers. **(B)** Radar chart showing the median scores of the four e-Rubric dimensions across the three feedback modalities, providing a multidimensional user experience profile. The radial axes represent the four dimensions: Usefulness (U), Ease of Use (EoU), Ease of Learning (EoL), and Satisfaction (S).

Despite the lack of statistical significance, the musical feedback showed descriptively higher median scores across all evaluated dimensions: Usefulness (4.50 vs. 4.12 for both visual and combined), Ease of Use (4.60 vs. 4.20 for visual and 4.30 for combined), Ease of Learning (4.00 vs. 3.67 for visual and 4.00 for combined), Satisfaction (4.72 vs. 4.17 for visual and 4.44 for combined), and Overall Usability (4.57 vs. 4.21 for visual and 4.24 for combined).

## Discussion

This study compared three biofeedback modalities (visual, musical and combined audiovisual), during intensity-guided stationary cycling in healthy adults. We aimed to address four research questions: (RQ1) What are the performance trade-offs between these modalities in terms of control strategy? (RQ2) Does combined feedback produce synergistic benefits or cognitive penalties? (RQ3) How do these modalities compare in usability and user satisfaction? (RQ4) Do these modalities maintain comparable physiological load?

Our findings reveal that the control variable encoded by biofeedback (cumulative position error versus instantaneous speed deviation) determines the control strategy adopted during intensity-guided exercise, with important implications for performance, physiological load, cognitive demands, and user experience.

### Performance and physiology: control strategy profiles (RQ1 & RQ4)

In response to RQ1, our data reveal a clear distinction between preventive and reactive control strategies. Position-based visual feedback (visual and combined conditions) promoted stable, preventive control characterized by high zone adherence (*>* 95% time in zone in Set 2), low rate of zone exits (*<* 2 exits/min), and minimal sustained deviations. This performance profile reflects a preventive control strategy where users continuously monitor their position relative to a moving target and make anticipatory corrections before errors (moving beyond the established limits) appear. The key advantage of position-based feedback lies in the provision of gradient information: users can perceive not only whether they are inside or outside the target limits, but also how far they deviate and the rate of change of this deviation. This control strategy enables feedforward control mechanisms [70–72], where users predict future states based on current trajectory and adjust proactively to prevent zone exits; and aligns with theoretical frameworks emphasizing the role of continuous sensory information in movement planning and online corrections [73–75].

Speed-based musical feedback showed a different performance profile. While maintaining good but slightly lower zone adherence overall (88–95% across sets), it produced approximately 10-fold higher rate of zone exits (14.52 exits/min in Set 1 vs. 1.48–1.79 for visual and combined, respectively) but enabled extremely rapid error corrections (∼0.2s recovery time vs. 1–3s for position-based systems). This pattern reflects a reactive control strategy driven by the threshold-based nature of the auditory cues. Unlike visual position feedback, musical tempo changes provide limited gradient information [57]: within the target zone, users receive no gradient information about proximity to boundaries, while outside the zone, progressive distortion in tempo and pitch indicates deviation magnitude. Users cannot anticipate threshold crossings before they occur; instead, they respond rapidly once the music indicates that a boundary has been crossed. The extremely rapid recovery times (∼0.2s) and the significantly reduced number of sustained deviations demonstrate the temporal precision advantages of auditory processing for time-dependent motor tasks [76–78]; while the high rate of target zone exits represents oscillation around the target threshold as opposed to sustained deviation from the target in one particular direction. This can be understood as a “hunting” behavior where rapid corrections maintain average speed near the prescribed target speed despite frequent boundary crossings [79, 80].

Therefore, the performance data reveal complementary strengths rather than a clear hierarchy between feedback modalities. Position-based visual feedback (visual and combined) optimizes for stability and precision, achieving the highest zone adherence with minimal rate of exits. This could be ideal for contexts requiring strict dosing or quantifiable adherence metrics (e.g., research protocols, outcome assessments). On the other hand, speed-based musical feedback optimizes for agility and ease of error correction, enabling rapid corrections and minimizing sustained deviations, which is quite relevant for populations for whom sustained compensatory efforts above the prescribed intensity might pose safety risks. Thus, neither strategy is inherently superior, but each serves distinct functional goals aligned with different clinical priorities, which we elaborate in our clinical translation recommendations.

Addressing our fourth research question regarding physiological load, it is important to note that these distinct control strategies did not result in distinct cardiovascular stress. Heart rate variability (*CV-HR*) showed no significant differences between groups (*p* = 0.85), indicating that the brief excursions above and below target speed limits in the musical condition did not promote a physiologically distinct cardiovascular load compared to the more stable trajectories in position-based conditions [81]. Thus, both preventive and reactive strategies were equally effective in maintaining the prescribed physiological dose.

However, position-based systems carry a previously underappreciated physiological cost: the compensatory effort penalty. When users fall behind the virtual target (due to accumulated positional error), they must generate sustained supra-target exertion to reduce the accumulated gap. This creates intermittent bursts of high-intensity effort that, while maintaining zone adherence metrics, might impose additional physical and cognitive demands beyond the prescribed exercise intensity. This has important implications for populations with cardiovascular comorbidities, where avoiding sustained supra-maximal exertion is often more important than preventing brief threshold crossings [82]. We return to this issue when discussing cognitive load below.

### Cognitive load: the multimodal integration paradox (RQ2)

Regarding our second research question, contrary to our hypothesis and theoretical predictions from multisensory integration research [48, 83], combining visual and musical modalities did not produce synergistic benefits. Instead, the combined condition incurred cognitive load penalties. The combined feedback condition produced significantly higher perceived physical demand compared to both musical (*p* = 0.009) and visual (*p* = 0.003) conditions, with median ratings approximately double those of musical feedback. The combined condition also required significantly higher effort than visual-only feedback (combined: 21.3 vs. visual: 14.0; *p* = 0.015). Consequently, the combined condition incurred the costs of position-based control without significantly improving overall adherence in Set 1; only in Set 2 did combined feedback showed improved overall adherence (% time in zone) over musical feedback (*p* = 0.008).

These findings likely reflect the compensatory effort penalty inherent to cummulative position-based feedback, understood through the distinction between error forgiveness and ease of error correction [84, 85]. While error forgiveness refers to the tolerance for deviation (which is designed to be equivalent in both modalities), ease of error correction describes the cost to return to the target state. Position-based systems act as an integral of error over time, creating a “positional debt” that forces users to sustain supra-target exertion to eliminate the gap (low ease of correction). In contrast, the musical condition functions as a speed-based mechanism with high ease of correction (∼0.2s recovery times). Once users adjust their pedaling speed, the auditory feedback instantly confirms the correction without requiring compensation for past deviations.

Our results suggest that redundant sensory coding does not automatically confer advantages in motor control tasks. Our combined feedback provided congruent position information through both visual and auditory channels, which matches the type of redundant coding that multisensory integration theory suggests should lead to improved efficiency [48, 86]. However, this redundancy imposed costs without commensurate performance benefits. This aligns with psychophysical evidence that visual and auditory signals do not always integrate automatically; as Burr and Alais [51] demonstrated, attentional resources are largely modality-specific, and redundant signals can increase attentional demands without improving perceptual resolution. In our case, the “more information is better” assumption was not fulfilled; the visual feedback’s demand to “pay back” positional debt likely overrode the possible musical benefit.

To address this, we propose that effective multimodal design for motor control may require orthogonal rather than redundant information, where each modality encodes distinct task dimensions, i.e. multimodal combination rather than integration [87, 88]. For cycling rehabilitation, this could involve visual feedback for bilateral pedaling symmetry [89] while musical feedback regulates pedaling speed, providing complementary streams. However, it is important to consider that orthogonal designs carry their own risks. Dividing attention between multiple information streams may increase cognitive load, particularly in populations with limited cognitive capacity. The optimal balance between redundant (potentially robust but cognitively costly) and orthogonal (potentially informative but attentionally demanding) multimodal designs [90] remains an open question requiring systematic investigation. An alternative approach might involve conditional activation where sensory channels activate selectively based on task state. For example, musical cues could activate only when error thresholds approach or when visual attention is directed elsewhere, or vice versa, providing complementary alerts rather than continuous redundant tracking. This could preserve the benefits of multimodal availability while avoiding the costs of continuous dual-channel processing.

These findings have direct implications for biofeedback design in rehabilitation contexts where minimizing compensatory effort is clinically important. For populations with limited exercise capacity, cardiovascular comorbidities, or fatigue-prone conditions, the high ease of error correction in speed-based feedback may promote safer and more sustainable training intensity [64, 91].

#### Usability and user satisfaction (RQ3)

Addressing our third research question, all three feedback modalities achieved high usability scores (*>* 4.0/5.0 across most dimensions), indicating strong feasibility and satisfaction. No statistically significant differences emerged across Usefulness, Ease of Use, Ease of Learning, Satisfaction, or Overall Usability dimensions (all *p >* 0.05).

However, musical feedback showed descriptively higher median scores across all evaluated dimensions, with Satisfaction ratings slightly elevated (musical: 4.72 vs. visual: 4.17 vs. combined: 4.44, though *p* = 0.089 n.s.). This trend toward higher satisfaction despite frequent zone exits (14.52 exits/min) suggests that ease of error correction observed in the speed-based feedback may be more important for user experience than ease of error prevention from the position-based modalities. This principle has particular relevance for neurological rehabilitation, where motor errors are inevitable and can trigger frustration spirals if attributed to functional impairment [92–95]. Position-based feedback systems naturally accumulate error information, creating scenarios where increasing effort is required to overcome accumulated deviations; a pattern that may undermine motivation and self-efficacy over repeated sessions [96]. Musical feedback’s immediate reset following speed corrections may help maintain consistent effort levels and positive emotional valence, avoiding the psychological burden of accumulated error. This error-forgiving design philosophy could promote sustained engagement better than error-preventing systems that maintain more strict control but demand sustained compensatory effort when deviations occur.

### Clinical translation recommendations

The different performance and cognitive profiles observed across feedback modalities suggest that optimal biofeedback design would depend on clinical priorities and patient characteristics.

For patient populations where motivation, long-term adherence, and cardiovascular safety are the most important concerns (e.g., home-based training, chronic rehabilitation, individuals with cardiovascular comorbidities or limited exercise capacity), musical-only feedback offers several advantages. The ease of error correction minimizes compensatory effort bursts while maintaining stable physiological training intensity. In addition, the immediate feedback reset following corrections may support positive emotional valence and sustained engagement across repeated training sessions. Rapid error corrections (∼0.2s) prevent sustained deviations that could exceed safety thresholds. It is also recommended and easy to include personalization features allowing selection of preferred music genres to enhance intrinsic motivation [53, 54, 97, 98].

For supervised clinical settings where precise dose control and objective adherence metrics are needed (e.g., acute rehabilitation, research protocols, outcome assessments), visual-only feedback provides stable control with moderate cognitive load, enabling precise quantification of adherence to prescribed intensity zones. The continuous gradient information about position relative to target supports development of feedforward motor planning abilities, as users learn to anticipate trajectory and adjust proactively before errors accumulate [70, 73]. In this case, clinicians should provide explicit instructions to avoid over-exertion and monitor for signs of excessive compensatory effort, particularly during longer training sessions. It is advisable to make use of the adjustable threshold strictness (already implemented in the system) allowing therapists to match each individual’s functional capacity and resistance to fatigue.

For multimodal systems, our findings suggest that future designs should prioritize conditional activation rather than simultaneous presentation. Musical cues could activate selectively when visual attention diverts or safety thresholds approach, providing complementary alerts rather than redundant tracking information. They could also incorporate orthogonal information mapping where each modality encodes distinct task dimensions (e.g., visual for bilateral pedaling symmetry [99], auditory for speed), while carefully evaluating the associated cognitive load. Finally, systems might benefit from adaptive weighting informed by individual preferences, cognitive capacity, and real-time performance [100–102], allowing the interface to emphasize the modality that is most effective for each user.

Beyond these context-specific guidelines, clinicians should assess individual preferences when selecting feedback modality. Prior experience with musical training or gaming, personal sensitivity to different sensory channels, and subjective comfort with each interface may significantly influence both immediate performance and long-term adherence. When clinical priorities allow flexibility, patient preference should be weighted alongside objective performance considerations.

### Limitations and future directions

This proof-of-concept study has several limitations that define important directions for future research. The single-session, non-counterbalanced design prevents assessment of learning effects or long-term adherence patterns. All participants performed the higher-intensity set before the lower-intensity set to mitigate fatigue and prioritize safety, but this confounds the isolation of learning effects and set-specific differences. The superior performance in Set 2 in most conditions may reflect familiarization with the control task rather than true differences in task intensity. Longitudinal validation studies with counterbalanced designs and multiple training sessions are needed to examine whether control strategies and preferences evolve over time, and whether initial satisfaction translates to sustained adherence and functional improvements.

Our findings in healthy young adults may not generalize to populations with motor impairments, reduced cognitive capacity, or cardiovascular limitations. Neurologically impaired populations may show different control strategy patterns, and the cognitive demands of multimodal feedback could overwhelm patients with attentional deficits or executive dysfunction. The physical demands and compensatory costs observed here could be amplified in individuals with reduced exercise capacity or fatigue-prone conditions. Randomized controlled trials in target clinical populations including stroke, spinal cord injury, and Parkinson’s disease should employ counterbalanced designs to isolate learning effects and assess functional outcomes, safety profiles, and long-term adherence. These trials must carefully document adverse events, dropout rates, and patient-reported outcomes to establish not only efficacy but also real-world feasibility and acceptability.

While the sample size was adequate for detecting the large effect sizes observed (*η*^2^ *>* 0.25 for key findings), the study was underpowered to detect small-to-medium effects and potential interactions between feedback modality and individual differences such as musical training, gaming experience, or motor learning capacity. Fixed intervention parameters including virtual environment aesthetics, threshold strictness, and musical tempo mapping were not systematically varied, which limits the ability to determine optimal parameter selection across diverse users. Systematic optimization studies should vary these parameters to identify optimal configurations for specific clinical contexts, enabling evidence-based customization rather than arbitrary design choices.

The difference in control strategies between position-based (visual and combined) and speed-based (musical) feedback creates inherent asymmetries that limit direct comparability. While our feedback-agnostic performance metrics enable quantitative comparison, the systems differ in how they transmit temporal and spatial information about task state. This represents a design feature reflecting genuine differences in how visual and auditory modalities provide information for motor control [57], rather than a methodological limitation. However, it complicates interpretation of relative performance and requires careful consideration of which performance aspects are therapeutically relevant for specific clinical contexts.

Finally, investigation of orthogonal multimodal designs where each sensory channel encodes different task dimensions (such as visual feedback for bilateral pedaling symmetry and auditory feedback for speed) should be performed, with careful assessment of cognitive load trade-offs. These designs may avoid the redundancy penalties observed in our combined condition while preserving multimodal benefits. Development of adaptive personalization algorithms that adjust feedback parameters based on real-time performance, user preferences, and cognitive and physical capacity represents another promising direction. Such systems could dynamically weight sensory modalities or adjust difficulty to match individual capabilities and maintain optimal challenge levels throughout rehabilitation.

## Conclusion

This study demonstrates that biofeedback modality determines the control strategy used during intensity-guided cycling rehabilitation. Visual-only (position-based) feedback promotes stable, preventive control with high adherence but imposes compensatory effort penalties; whereas musical-only (speed-based) feedback enables agile, reactive control with rapid error corrections and lower perceived effort. Combining these modalities did not improve performance; instead, redundant integration incurred cognitive costs without performance advantages, challenging the assumption that multimodal feedback is inherently superior.

Our findings suggest that the ease of error correction and the rapid return to positive feedback state (observed in the musical feedback group) may be more significant for user satisfaction and sustained engagement than ease of error prevention. This is particularly relevant for rehabilitation contexts, where motor errors may be inevitable and preserving motivation is essential for therapy adherence.

This work moves beyond “one-size-fits-all” approaches toward an evidence-based framework for personalized exercise prescription. By identifying the trade-offs between control stability, physiological safety, cognitive load, and user experience, rehabilitation systems can be tailored to match specific clinical goals. Musical feedback is recommended for home-based training emphasizing adherence and safety; visual feedback for supervised settings requiring precise dose control and feedforward motor planning development; and future development must focus on orthogonal information mapping or conditional activation to explore the true potential of multimodal biofeedback.

## Supporting information

**S1 Fig.**
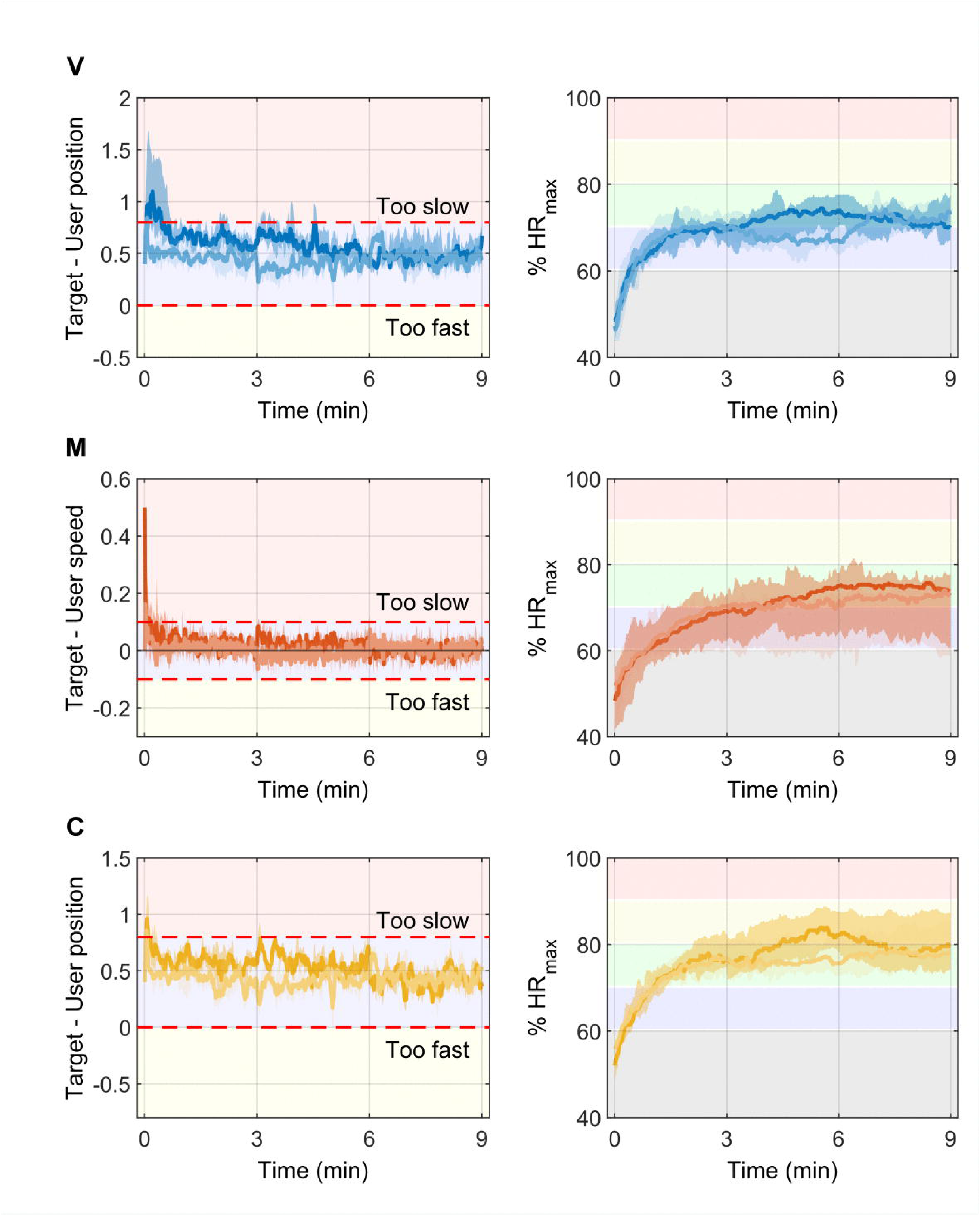
Median pedaling speed deviation relative to target zone boundaries and heart rate trajectories during exercise sets. (**A**) Time-series analysis of difference between target pedaling position/speed and actual user position/speed for visual-only (V, top), musical-only (M, middle), and combined audiovisual (C, bottom) conditions for both Set 1 (darker) and Set 2 (lighter). The colored rectangular zones indicate different levels of deviation from target position or speed: reddish indicates “too slow” (below target), blueish indicates “within target range,” and yellowish indicates “too fast” (above target). The dashed red lines represent the boundaries between zones. Solid lines represent median trajectories and shaded regions represent interquartile range (25th to 75th percentiles) across participants in each condition. (**B**) Heart rate as a percentage of age-predicted maximum heart rate (HRmax) for both Set 1 (darker) and Set 2 (lighter), with colored horizontal bands representing intensity zones: light blue (*<*60% HRmax, light intensity), green (60-70% HRmax, moderate intensity), yellow (70-85% HRmax, vigorous intensity), and pink (*>*85% HRmax, near-maximal intensity). Solid lines represent median trajectories and shaded regions represent interquartile range (25th to 75th percentiles) across participants in each condition. These trajectories confirm that all three feedback modalities successfully maintained participants within the prescribed moderate-to-high cardiovascular training zones (70% HRmax baseline) despite employing distinct control strategies.

**S2 Fig.**
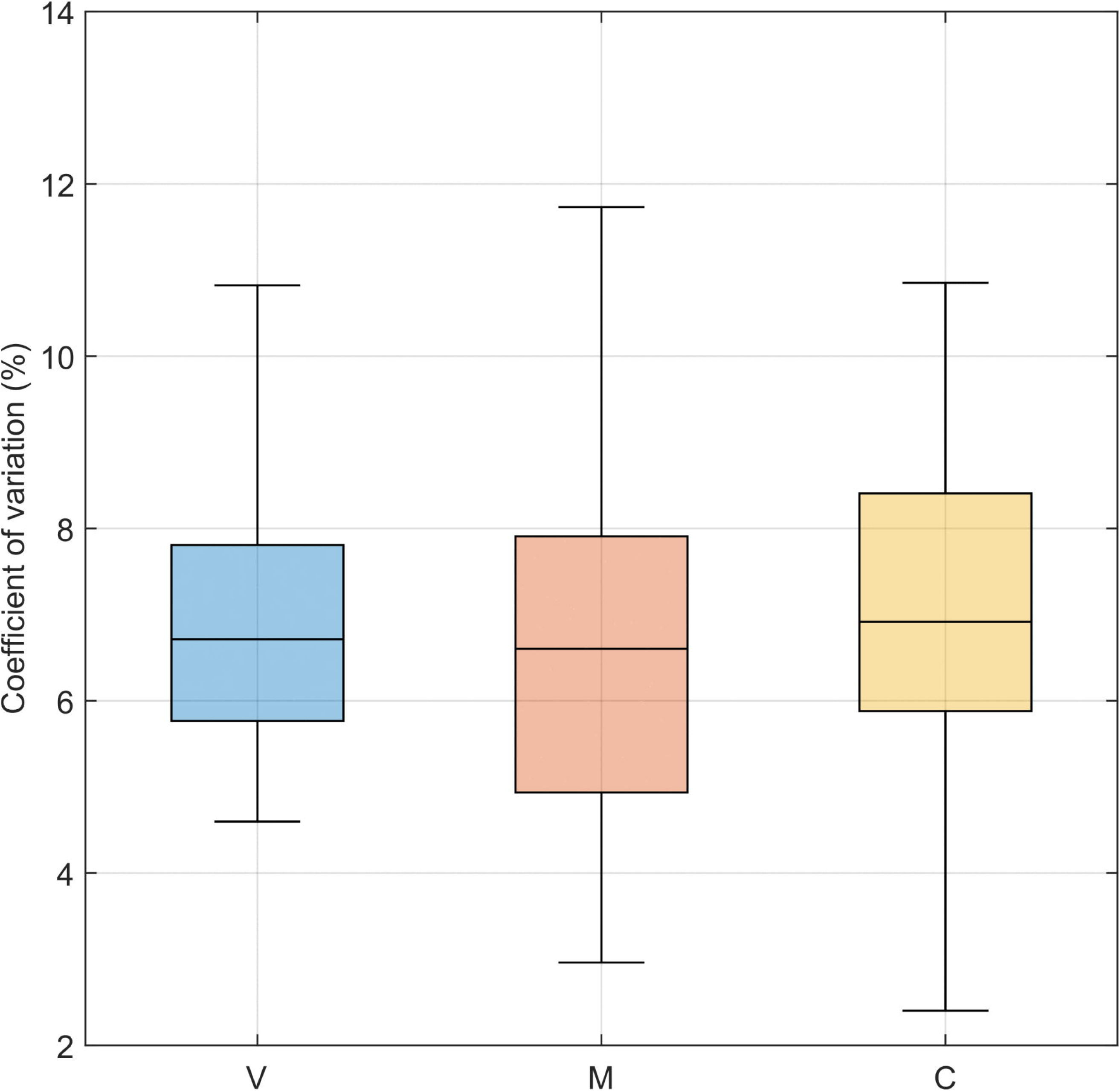
Heart rate variability across feedback modalities. Distribution of the coefficient of variation of heart rate (CV-HR, expressed as percentage) for visual-only (V, blue), musical-only (M, orange), and combined audiovisual (C, yellow) feedback conditions. The red line within each box indicates the median coefficient of variation, the box edges represent the interquartile range (25th to 75th percentiles), and the whiskers extend to the most extreme data points not considered outliers. No significant differences were observed between conditions (ANOVA: *p* = 0.8518, *η*^2^ = 0.006), confirming equivalent cardiovascular training intensity despite different control strategies.

**S3 Fig.**
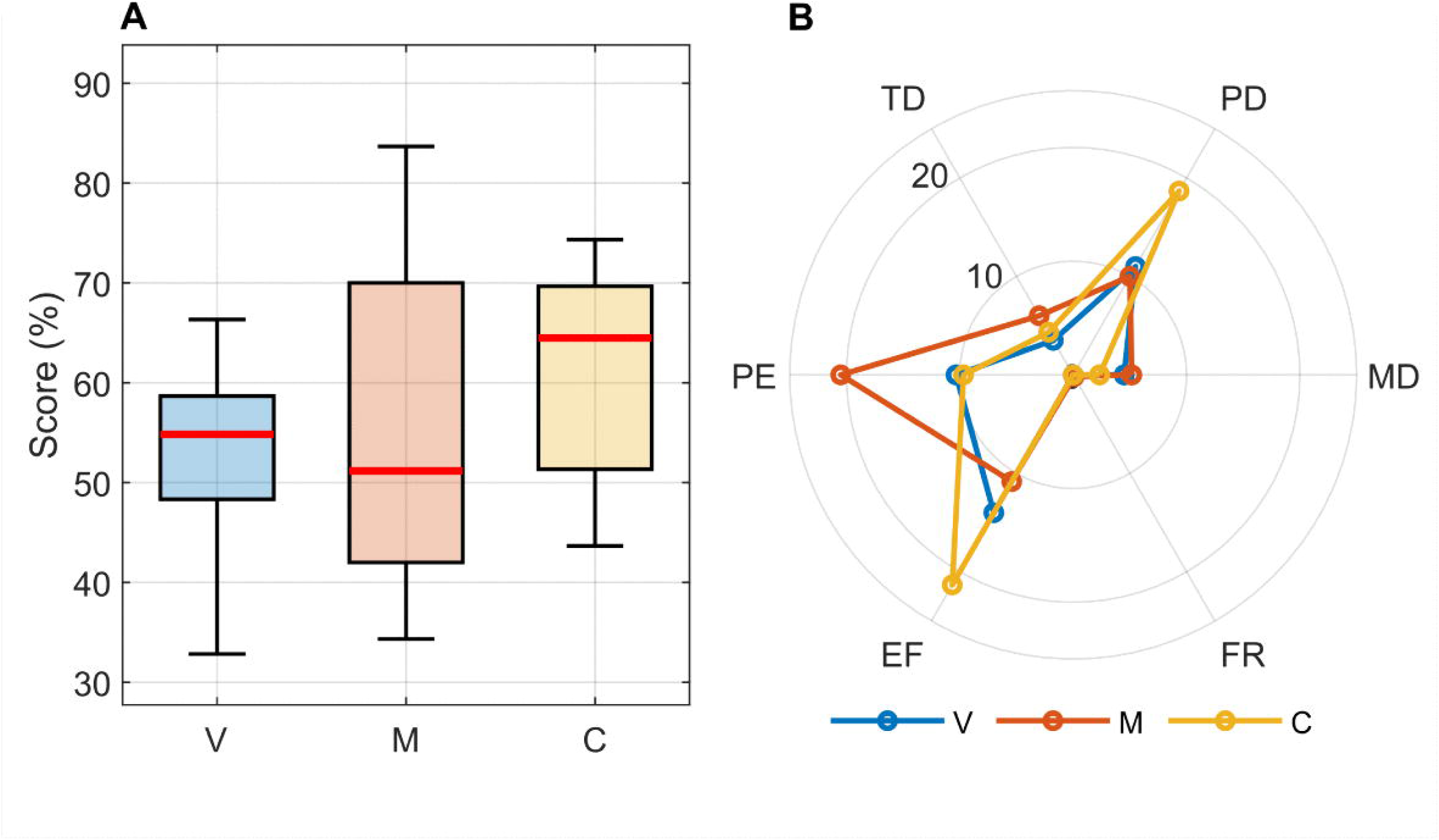
Overall cognitive workload and multidimensional profile. **(A)** Distribution of weighted overall NASA-TLX scores (0–100% scale) across visual-only (V, blue), musical-only (M, orange), and combined audiovisual (C, yellow) feedback conditions. The red line within each box indicates the median overall workload score, the box edges represent the interquartile range (25th to 75th percentiles), and the whiskers extend to the most extreme reported scores. No significant differences in overall cognitive workload were detected between conditions (ANOVA: *p* = 0.2450). **(B)** Radar chart illustrating the mean scores for the six NASA-TLX subscales across the three feedback modalities, providing a multidimensional cognitive workload profile. The radial axes represent the six dimensions and each colored line connects the median scores for one feedback condition, revealing the workload profile characteristic of each modality. **(MD)** Mental Demand. **(PD)** Physical Demand. **(TD)** Temporal Demand. **(PE)** Performance. **(EF)** Effort. **(FR)** Frustration.

**S4 Fig.**
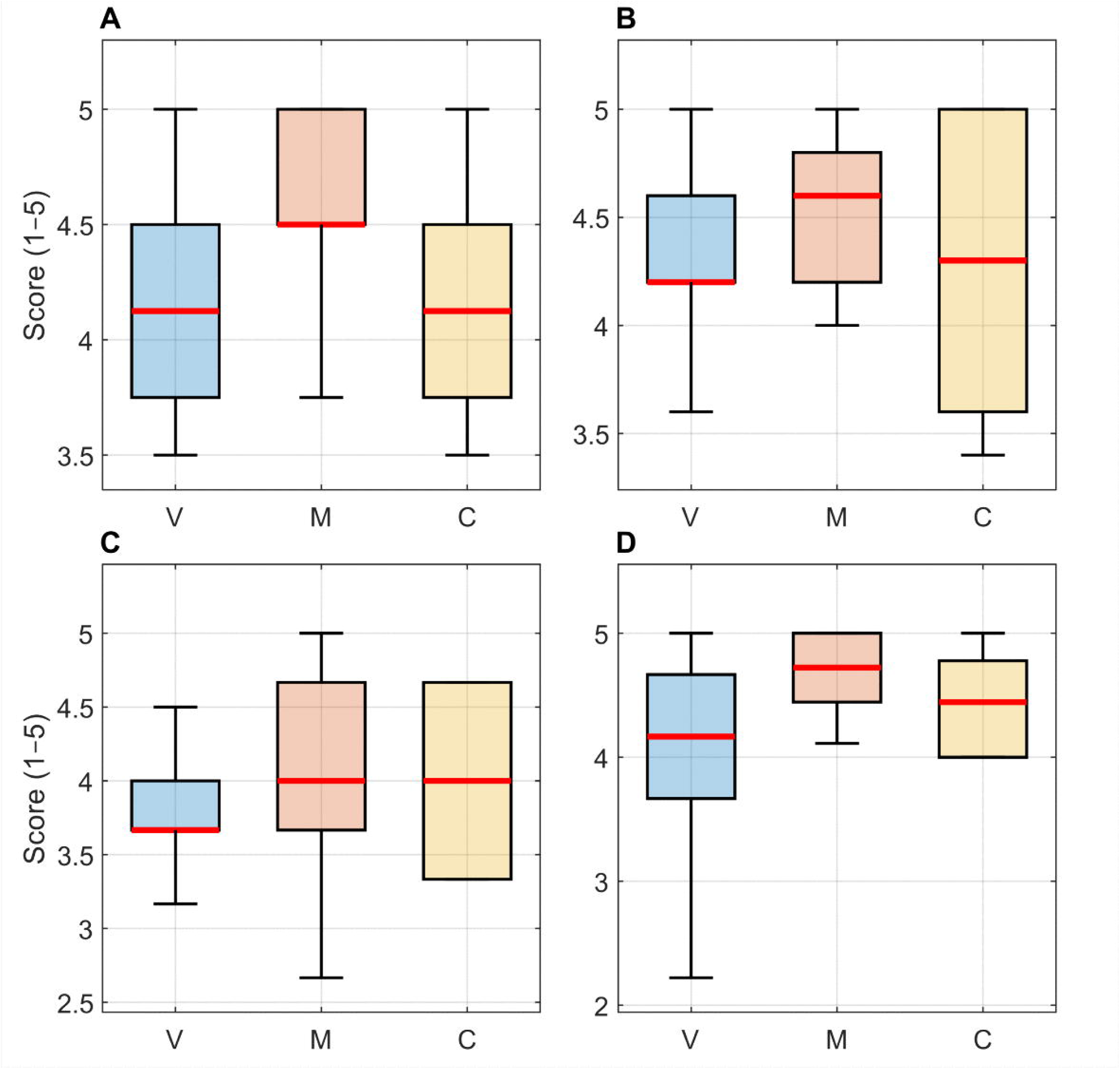
User experience scores across usability dimensions. Distribution of user experience and usability scores (on a 1 to 5 Likert scale) reported using the e-Rubric questionnaire for the three feedback modalities: visual-only (V, blue), musical-only (M, orange), and combined audiovisual (C, yellow). The red line within each box indicates the median score, the box edges represent the interquartile range (25*^th^* to 75*^th^* percentiles), and the whiskers extend to the most extreme reported scores not considered outliers. **(A)** Usefulness. **(B)** Ease of use. **(C)** Ease of learning. **(D)** Satisfaction.

**S1 Table.** Performance metrics for Set 1 by feedback modality: Descriptive statistics. Comprehensive performance measures during the first 9-minute exercise session (Set 1: 3 min baseline, 3 min +15% intensity, 3 min baseline) for visual-only, musical-only, and combined audiovisual feedback conditions. Values are presented as median [IQR]. Metrics include: percentage of time spent within target speed zone (% time in zone), rate of exits from target zone (exits/min), median recovery time following zone exits (seconds), and number of sustained deviations (periods *>*5 consecutive seconds outside target zone).

| Metric | Visual | Musical | Combined |
| --- | --- | --- | --- |
| % Time in zone | 89.42 [80.41–95.88] | 88.90 [86.55–92.75] | 95.75 [92.10–97.60] |
| Exits/min | 1.48 [1.00–3.05] | 14.52 [11.19–23.07] | 1.79 [1.31–2.18] |
| Recovery time (s) | 2.48 [0.87–3.84] | 0.21 [0.19–0.25] | 1.06 [0.92–1.43] |
| Sustained deviations | 2.50 [1.00–4.00] | 0.50 [0.00–1.00] | 0.50 [0.00–3.00] |

Values shown as Median [Q1–Q3]. See S2 Table for statistical comparisons.

**S2 Table.** Statistical analysis for Set 1 performance metrics. Statistical comparisons across feedback modalities for performance measures during Set 1. Test selection was based on data distribution and variance homogeneity. Effect sizes (*η*^2^) are interpreted as small (≥0.01), medium (≥0.06), and large (≥0.14). Post-hoc pairwise comparisons were conducted with Bonferroni-adjusted significance threshold (*α* = 0.0167).

| Metric | Test | p-value | $\eta^2$ | Post-hoc (p-corrected) |
| --- | --- | --- | --- | --- |
| % Time in zone | Welch ANOVA | 0.0335* | 0.155 | — |
| Exits/min | Welch ANOVA | 0.0006** | 0.661 | M-V (0.0005), M-C (0.0005) |
| Recovery time (s) | Welch ANOVA | <0.0001** | 0.479 | M-V (0.0022), M-C (0.0001) |
| Sustained deviations | Kruskal-Wallis | 0.0422* | 0.173 | M-V (0.0109) |

\* $p < 0.05$ , \*\* $p < 0.01$ . Post-hoc: Games-Howell or Mann-Whitney ( $\alpha = 0.0167$ ). V = Visual, M = Musical, C = Combined.

**S3 Table.** Performance metrics for Set 2 by feedback modality: Descriptive statistics. Comprehensive performance measures during the second 9-minute exercise session (Set 2: 3 min baseline, 3 min -15% intensity, 3 min baseline) for visual-only, musical-only, and combined audiovisual feedback conditions. Values are presented as median [IQR]. Metrics include: percentage of time spent within target speed zone (% time in zone), rate of exits from target zone (exits/min), median recovery time following zone exits (seconds), and number of sustained deviations (periods *>*5 consecutive seconds outside target zone).

| Metric | Visual | Musical | Combined |
| --- | --- | --- | --- |
| % Time in zone | 97.28 [87.79–99.81] | 95.17 [86.26–95.99] | 98.13 [97.10–99.48] |
| Exits/min | 0.38 [0.11–1.84] | 9.41 [7.19–21.67] | 0.54 [0.32–0.55] |
| Recovery time (s) | 1.60 [0.94–2.77] | 0.22 [0.21–0.23] | 1.80 [0.66–2.21] |
| Sustained deviations | 1.00 [0.00–4.00] | 0.00 [0.00–0.00] | 0.50 [0.00–1.00] |

Values shown as Median [Q1–Q3]. See S4 Table for statistical comparisons.

**S4 Table.** Statistical analysis for Set 2 performance metrics. Statistical comparisons across feedback modalities for performance measures during Set 2. Test selection was based on data distribution and variance homogeneity. Effect sizes (*η*^2^) are interpreted as small (≥0.01), medium (≥0.06), and large (≥0.14). Post-hoc pairwise comparisons were conducted with Bonferroni-adjusted significance threshold (*α* = 0.0167).

| Metric | Test | p-value | $\eta^2$ | Post-hoc (p-corrected) |
| --- | --- | --- | --- | --- |
| % Time in zone | Welch ANOVA | 0.0164* | 0.190 | M-C (0.0077) |
| Exits/min | Kruskal-Wallis | 0.0001** | 0.573 | M-V (0.0002), M-C (0.0002) |
| Recovery time (s) | Welch ANOVA | 0.0009** | 0.266 | M-C (0.0014) |
| Sustained deviations | Kruskal-Wallis | 0.0369* | 0.249 | — |

\* $p < 0.05$ , \*\* $p < 0.01$ . Post-hoc: Games-Howell or Mann-Whitney ( $\alpha = 0.0167$ ). V = Visual, M = Musical, C = Combined.

**S5 Table.** NASA-TLX cognitive workload scores by feedback modality: Descriptive statistics. Weighted dimension scores (0–100% scale) for the six subscales of the NASA Task Load Index and the overall weighted workload score across visual-only, musical-only, and combined audiovisual feedback conditions. Values are presented as median [IQR]. Dimensions assessed include: Mental Demand (MD), Physical Demand (PD), Temporal Demand (TD), Performance (PE), Effort (EF), and Frustration (FR). Higher scores indicate greater workload for all dimensions except Performance, where higher scores indicate better perceived performance.

| Dimension | Visual | Musical | Combined |
| --- | --- | --- | --- |
| MD | 4.5 [1.3–11.0] | 5.2 [2.7–6.7] | 2.3 [0.7–4.0] |
| PD | 11.0 [7.0–14.7] | 10.0 [1.3–14.0] | 18.7 [16.0–26.7] |
| TD | 3.5 [1.0–10.0] | 6.0 [0.7–12.0] | 4.3 [3.3–10.0] |
| PE | 10.3 [6.0–14.0] | 20.5 [6.7–30.0] | 9.7 [9.3–17.0] |
| EF | 14.0 [5.0–23.3] | 10.8 [6.7–12.0] | 21.3 [18.3–23.3] |
| FR | 0.0 [0.0–0.0] | 0.2 [0.0–2.0] | 0.0 [0.0–0.0] |
| TLX<sub>total</sub> | 54.8 [48.3–58.7] | 51.2 [42.0–70.0] | 64.5 [51.3–69.7] |

Values shown as Median [Q1–Q3]. See S6 Table for statistical comparisons. *Abbreviations:* MD, Mental Demand; PD, Physical Demand; TD, Temporal Demand; PE, Performance; EF, Effort; FR, Frustration.

**S6 Table.** Statistical analysis for NASA-TLX dimensions. Statistical comparisons across feedback modalities for cognitive workload dimensions. Test selection was based on data distribution and variance homogeneity. Effect sizes (*η*^2^) are interpreted as small (≥0.01), medium (≥0.06), and large (≥0.14). Post-hoc pairwise comparisons were conducted with Bonferroni-adjusted significance threshold (*α* = 0.0167). Combined feedback showed significantly higher physical demand compared to both musical and visual conditions, and significantly higher effort compared to visual-only feedback.

| Dimension | Test | p-value | $\eta^2$ | Post-hoc (p-corrected) |
| --- | --- | --- | --- | --- |
| MD | Kruskal-Wallis | 0.1486 | 0.108 | — |
| PD | ANOVA | 0.0016** | 0.378 | M-C (0.0090), V-C (0.0025) |
| TD | ANOVA | 0.8039 | 0.016 | — |
| PE | ANOVA | 0.1164 | 0.147 | — |
| EF | ANOVA | 0.0189* | 0.255 | V-C (0.0147) |
| FR | Kruskal-Wallis | 0.1725 | 0.098 | — |
| TLX<sub>total</sub> | ANOVA | 0.2450 | 0.099 | — |

\* $p < 0.05$ , \*\* $p < 0.01$ . Post-hoc: Tukey-Kramer or Mann-Whitney ( $\alpha = 0.0167$ ). V = Visual, M = Musical, C = Combined. *Abbreviations*: MD, Mental Demand; PD, Physical Demand; TD, Temporal Demand; PE, Performance; EF, Effort; FR, Frustration.

**S7 Table.** User experience and usability scores by feedback modality: Descriptive statistics. Scores (1–5 Likert scale) from the e-Rubric usability questionnaire across visual-only, musical-only, and combined audiovisual feedback conditions. Values are presented as median [IQR]. Dimensions evaluated include: Usefulness, Ease of Use, Ease of Learning, Satisfaction and Overall Usability. All three feedback modalities achieved high usability scores across dimensions, with median scores generally exceeding 4.0 on the 5-point scale.

| Dimension | Visual | Musical | Combined |
| --- | --- | --- | --- |
| Usefulness | 4.25 [3.50–4.50] | 4.50 [4.50–5.00] | 4.50 [4.00–4.50] |
| Ease of Use | 4.21 [4.00–4.43] | 4.64 [4.43–4.86] | 4.36 [4.14–4.71] |
| Ease of Learning | 4.00 [3.25–4.25] | 4.25 [3.25–4.75] | 4.12 [3.50–4.25] |
| Satisfaction | 4.38 [3.50–4.75] | 4.75 [4.50–5.00] | 4.25 [4.00–4.75] |
| Overall Usability | 4.21 [3.76–4.38] | 4.57 [4.29–4.62] | 4.24 [3.95–4.62] |

Values shown as Median [Q1–Q3]. All scores rated on 1–5 Likert scale. See S8 Table for statistical comparisons.

**S8 Table.** Statistical analysis for usability dimensions. Statistical comparisons across feedback modalities for user experience and usability dimensions assessed through the e-Rubric questionnaire. Test selection was based on data distribution and variance homogeneity. Effect sizes (*η*^2^) are interpreted as small (≥0.01), medium (≥0.06), and large (≥0.14). Post-hoc pairwise comparisons were conducted with Bonferroni-adjusted significance threshold (*α* = 0.0167). Although Satisfaction showed a significant global effect, no pairwise comparisons reached significance after correction.

| Dimension | Test | p-value | $\eta^2$ | Post-hoc (p-corrected) |
| --- | --- | --- | --- | --- |
| Usefulness | Kruskal-Wallis | 0.1400 | 0.123 | — |
| Ease of Use | ANOVA | 0.0995 | 0.157 | — |
| Ease of Learning | ANOVA | 0.8570 | 0.011 | — |
| Satisfaction | Welch ANOVA | 0.0418* | 0.169 | — |
| Overall Usability | ANOVA | 0.0698 | 0.179 | — |

\* $p < 0.05$ . Post-hoc: Games-Howell ( $\alpha = 0.0167$ ). V = Visual, M = Musical, C = Combined. Although Satisfaction showed a significant global effect ( $p = 0.0418$ ), no pairwise comparisons reached significance after correction (M vs V: $p = 0.0583$ , M vs C: $p = 0.0411$ ).

**S1 Appendix. Complete e-Rubric usability questionnaire.** The questionnaire was used to evaluate usability and user experience. It was administered in Spanish, the participants’ native language. The English translations below are provided for reference. The questionnaire comprises 21 applicable items rated on a 5-point Likert scale, grouped into four dimensions following the USE framework: Usefulness, Ease of Use, Ease of Learning, and Satisfaction. Items originally worded negatively were reverse-scored during analysis so that higher scores consistently indicate better usability across all items. Notice that item 6 was removed since it was unrelated.

| Item No. | Question (English translation) | Dimension |
| --- | --- | --- |
| 1 | It was easy to use, without great effort. | Ease of Use |
| 2 | Incompatibilities appeared during use that made handling difficult. | Ease of Use |
| 3 | The environment was pleasant. | Satisfaction |
| 4 | The tool was exhausting and frustrating. | Ease of Use |
| 5 | It can be used without the need for prior explanations. | Ease of Learning |
| 6 | It was problematic to save and display your work. | Removed |
| 7 | It was motivating. | Satisfaction |
| 8 | I would prefer to have used another known tool instead of this one. | Satisfaction |
| 9 | It was simple, without unnecessary additions or complications. | Ease of Use |
| 10 | I felt insecure because the tool could be damaged by what I was doing. | Usefulness |
| 11 | I found the expected functions well integrated. | Satisfaction |
| 12 | The tool does not encourage its use. | Satisfaction |
| 13 | The tool can be understood very quickly by most users. | Ease of Learning |
| 14 | I encountered technical problems. | Usefulness |
| 15 | It performs the functions adequately to my expectations. | Usefulness |
| 16 | It takes more time with this tool than with another. | Usefulness |
| 17 | In case of need, I would use the tool again in the future. | Satisfaction |
| 18 | The language used was difficult to understand to interact with the tool. | Ease of Learning |
| 19 | It requires the help of a computer technician. | Ease of Use |
| 20 | I disagree with the graphical design of the tool. | Satisfaction |
| 21 | I would recommend this tool to other users. | Satisfaction |
| 22 | Overall, I am satisfied with the tool. | Satisfaction |

**S1 Video. Demonstration of the combined audiovisual feedback system.** Video recording illustrating the integrated virtual reality environment and musical biofeedback during stationary cycling. The video shows the first-person perspective visible to participants, including the virtual track, target rabbit, and environmental cues (sky color changes), synchronized with real-time musical tempo modulation corresponding to pedaling speed deviations. This demonstration provides visual and auditory context for understanding the multimodal feedback implementation described in the Methods section.

## Data Availability

The datasets used and/or analyzed during the current study are available from the corresponding author on reasonable request

https://zenodo.org/records/10925273

## Acknowledgments

The authors would like to thank all the volunteer participants that have contributed to this study.

